# BRASH syndrome as a clinical syndrome driven by polypharmacy: a pharmacovigilance study of 1,081 cases from FAERS

**DOI:** 10.64898/2026.01.15.26344203

**Authors:** Ilaria Costantini, Andrea Breglia, Giovanni Mantelli, Mirko Zanatta, Giorgio Ricci

## Abstract

**Background:** BRASH syndrome, characterized by bradycardia, renal failure, atrioventricular nodal blockade, shock, and hyperkalemia, is a recently described clinical entity that remains underrecognized. Current evidence is limited to case reports and small case series, and the pharmacological patterns underlying this syndrome have not been systematically evaluated.

**Methods:** We conducted a retrospective pharmacovigilance study using the U.S. Food and Drug Administration Adverse Event Reporting System (FAERS) from January 2004 to March 2025. Individual case safety reports explicitly coded as BRASH syndrome were identified and deduplicated. Associations between BRASH syndrome and individual drugs were assessed using Reporting Odds Ratios (RORs) and Information Components (ICs) with 95% confidence intervals. Drugs showing positive disproportionality signals were categorized according to their presumed role in the BRASH pathophysiological cascade.

**Results:** A total of 1,081 reports were included. The median age of patients was 74 years (interquartile range, 64–82); 45.5% were female, 38.7% were male, and sex was not reported in 15.8% of cases. All cases were classified as serious adverse drug reactions, including 7.1% fatal and 34.4% life-threatening events. Strong disproportionality signals were observed for atrioventricular nodal blocking agents, including metoprolol (ROR, 58.6; 95% CI, 52.0–66.1), verapamil (ROR, 52.5; 95% CI, 43.4–63.3), and carvedilol (ROR, 28.7; 95% CI, 24.4–33.6). Additional signals involved drugs plausibly contributing to renal dysfunction or hyperkalemia. Multiple pharmacologically relevant drugs were frequently reported within individual cases.

**Conclusions:** BRASH syndrome is associated with combinations of cardiovascular and renal-active medications rather than single-drug exposure, supporting its interpretation as a polypharmacy-driven clinical syndrome in vulnerable patients.

## INTRODUCTION

The BRASH syndrome, an acronym for Bradycardia, Renal failure, Atrioventricular nodal blockade, Shock, and Hyperkalemia, is a recently recognized and potentially fatal clinical entity. It has been described as a self-perpetuating vicious cycle, typically occurring in patients, often elderly, with underlying cardiac and renal impairment, even if mild, who are treated with atrioventricular (AV) nodal blocking agents such as beta-blockers or calcium channel blockers (1).

The syndrome is usually triggered by a relatively mild insult, such as dehydration or a gastrointestinal illness, leading to acute kidney injury and subsequent hyperkalemia. An increase in serum potassium level, even if moderate, synergizes with the pharmacological effects of AV nodal blocking medications, which may also accumulate due to impaired renal clearance. The result is disproportionately severe bradycardia and hypotension, which may lead to to shock. The ensuing reduction in cardiac output further compromises renal perfusion, thereby exacerbating acute kidney injury and worsening hyperkalemia, ultimately reinforcing and accelerating this vicious cycle.

Although BRASH syndrome was first described in 2016, it remains a critical diagnosis to consider in clinical practice. Its presentation can be heterogeneous, and the associated bradycardia is often refractory to standard chronotropic agents, an important feature distinguishing it from isolated hyperkalemia or pure drug overdose. Early recognition and prompt, simultaneous treatment of all components of the syndrome, including stabilization of cardiac rhythm, correction of hyperkalemia, and restoration of renal perfusion, are essential to interrupt the cycle and reduce the high associated morbidity and mortality. Despite its clinical relevance, BRASH syndrome remains widely underrecognized, underscoring the need for a higher index of suspicion among clinicians (2).

Since its initial description, numerous case reports have highlighted the heterogeneity of clinical patterns, triggers, and presentations of BRASH syndrome. However, at the moment, there are no large epidemiological studies that have systematically evaluated which drug classes or specific medications are most commonly associated with this condition. In addition, the overall pharmacological burden, intended as the number of concomitant medications implicated in the development of BRASH syndrome, has not been adequately explored.

To address this gap, we conducted an analysis of adverse event reportings submitted to the U.S. Food and Drug Administration Adverse Event Reporting System (FAERS). The aims of this pharmacovigilance study is to identify the main active substances among atrioventricular nodal blocking agents and other medications involved in BRASH syndrome, to characterize the patients most frequently affected in terms of age and sex, and to evaluate the pharmacological burden by assessing the association between this condition and the frequency of reported drugs.

## METHODS

A pharmacovigilance analysis was conducted using data from the U.S. Food and Drug Administration Adverse Event Reporting System (FAERS). Data were processed in the R statistical environment (R Foundation for Statistical Computing, Vienna, Austria) using the DiAna package (Disproportionality Analysis tool), which provides access to cleaned, deduplicated, and standardized FAERS datasets. Reports were retrieved from FAERS from its inception (from January 1, 2004 to March 31, 2025(3).

Adverse events were coded according to the Medical Dictionary for Regulatory Activities (MedDRA), and cases were identified using the Preferred Term (PT) “*brash syndrome*”. Extracted variables included patient demographics (age, sex, country), reporting characteristics (date of report), clinical outcomes, primary and secondary suspect drugs, and interacting drugs. For each report, the number of suspect, interacting, and concomitant drugs was recorded, and mean values were calculated for each category.

Drug analyses were conducted at the substance level. Only drugs reported as primary suspect, secondary suspect, or interacting were included in the analyses.

Descriptive statistics were used to summarize the data: categorical variables were reported as frequencies and percentages, while continuous variables were expressed as mean (± standard deviation, SD) or median (interquartile range, IQR), as appropriate. Disproportionality analyses were performed to assess the association between BRASH syndrome and individual drugs or drug classes by calculating Reporting Odds Ratios (RORs) and Information Components (ICs) with corresponding 95% confidence intervals. Disproportionality analyses are hypothesis-generating methods used in spontaneous reporting systems to detect safety signals by comparing the observed reporting frequency of a specific drug–adverse event combination with the expected frequency based on the overall database. These measures do not estimate incidence, relative risk, or causality, but identify signals of disproportionate reporting warranting further investigation.

The Reporting Odds Ratio (ROR) is *a* frequentist measure of disproportionality and was calculated as **ROR = (a/c) / (b/d)**, where a represents the number of reports of the drug of interest with the adverse event of interest, *b* the number of reports of the same drug with other adverse events, *c* the number of reports of the adverse event with other drugs, and *d* all remaining reports in the database. The **Information Component (IC)** is a Bayesian measure of disproportionality that compares observed and expected reporting and was calculated as **IC = log**_**2**_ **[a·(a + b + c + d) / ((a + b)(a + c))]**.

A **signal of disproportionate reporting** was defined by the presence of **at least three reports** for a given drug–event combination and a **lower bound of the 95% credibility interval (IC025) greater than zero** for the IC or a **lower bound of the 95% confidence interval greater than 1** for the ROR

Drugs showing a positive signal of disproportionate reporting were subsequently classified according to their presumed pharmacological and clinical role into four categories by two authors:

i. atrioventricular nodal blocking agents;
ii. drugs plausibily involved in the BRASH vicious circle, contributing to renal dysfunction, electrolyte imbalance, or hemodynamic instability;
iii. drugs associated with BRASH syndrome but likely reflecting therapeutic management of the condition rather than etiological involvement; and
iv. drugs reported in association with BRASH syndrome without an evident pathophysiological role.

This classification was performed to support clinical interpretation of disproportionality findings.

## RESULTS

By March 2025, a total of 1,081 deduplicated individual case safety reports (ICSRs) in which BRASH syndrome was reported as an adverse drug reaction (ADR) were retrieved from FAERS. Female patients accounted for 45.5% of cases, while sex was not reported in approximately 16% of reports. The median age was 74 years (interquartile range [IQR] 64–82). (male? Visto che si parla di un 16% senza indicazione del sesso?)

The first BRASH syndrome report was submitted in 2019, followed by a progressive increase in reporting over subsequent years. The geographical distribution of reports is shown in **Figure 1**, with the United States accounting for the majority of cases (n = 846; 78.3%).

**Figure 1.**
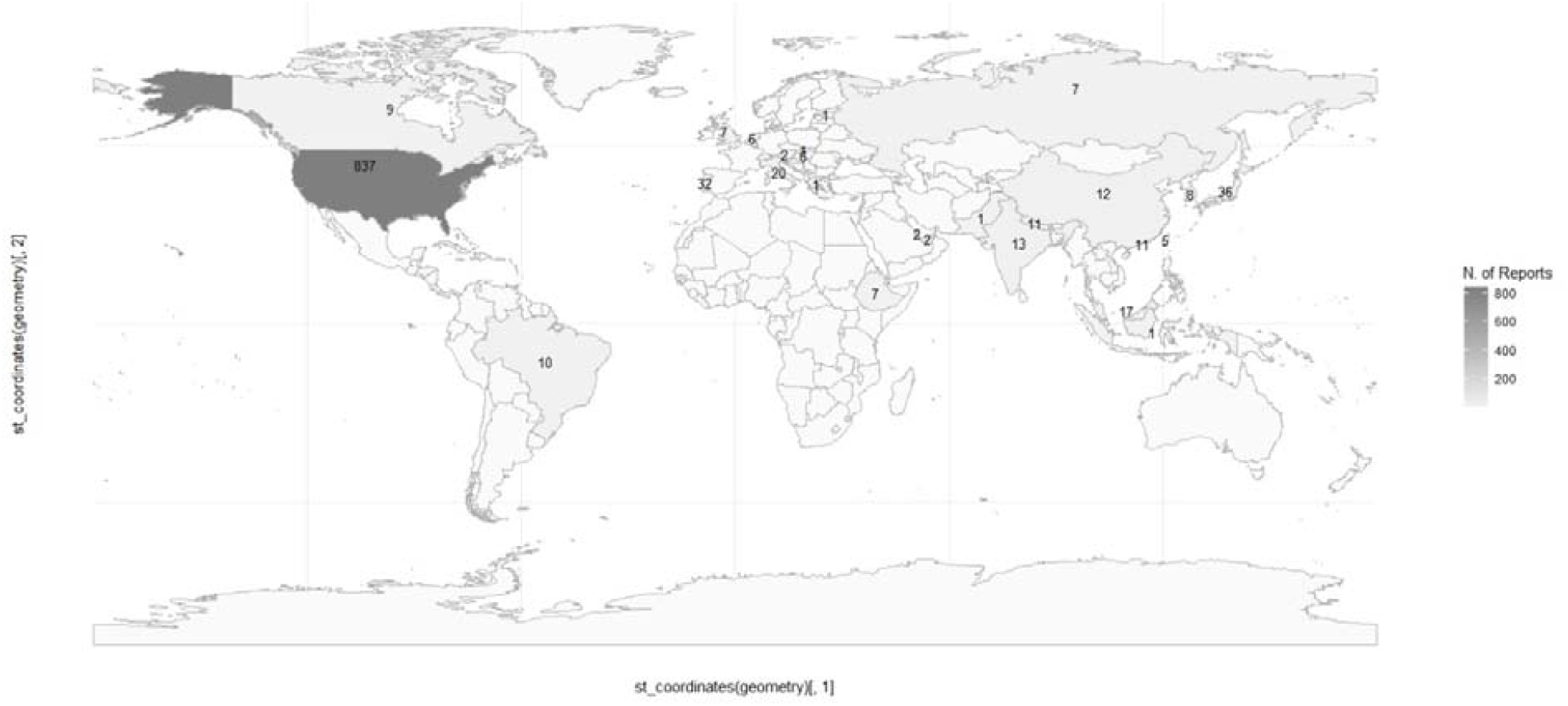
Geographical distribution of BRASH syndrome reports collected in FAERS (N=1,081). Shading intensity indicates the number of reports per country reports with darker areas corresponding to higher counts.

All reports were classified as serious ADRs (n = 1,081; 100%), including 77 deaths (7.1%) and 372 life-threatening events (34.4%) (**Table 1**).

**Table 1.**
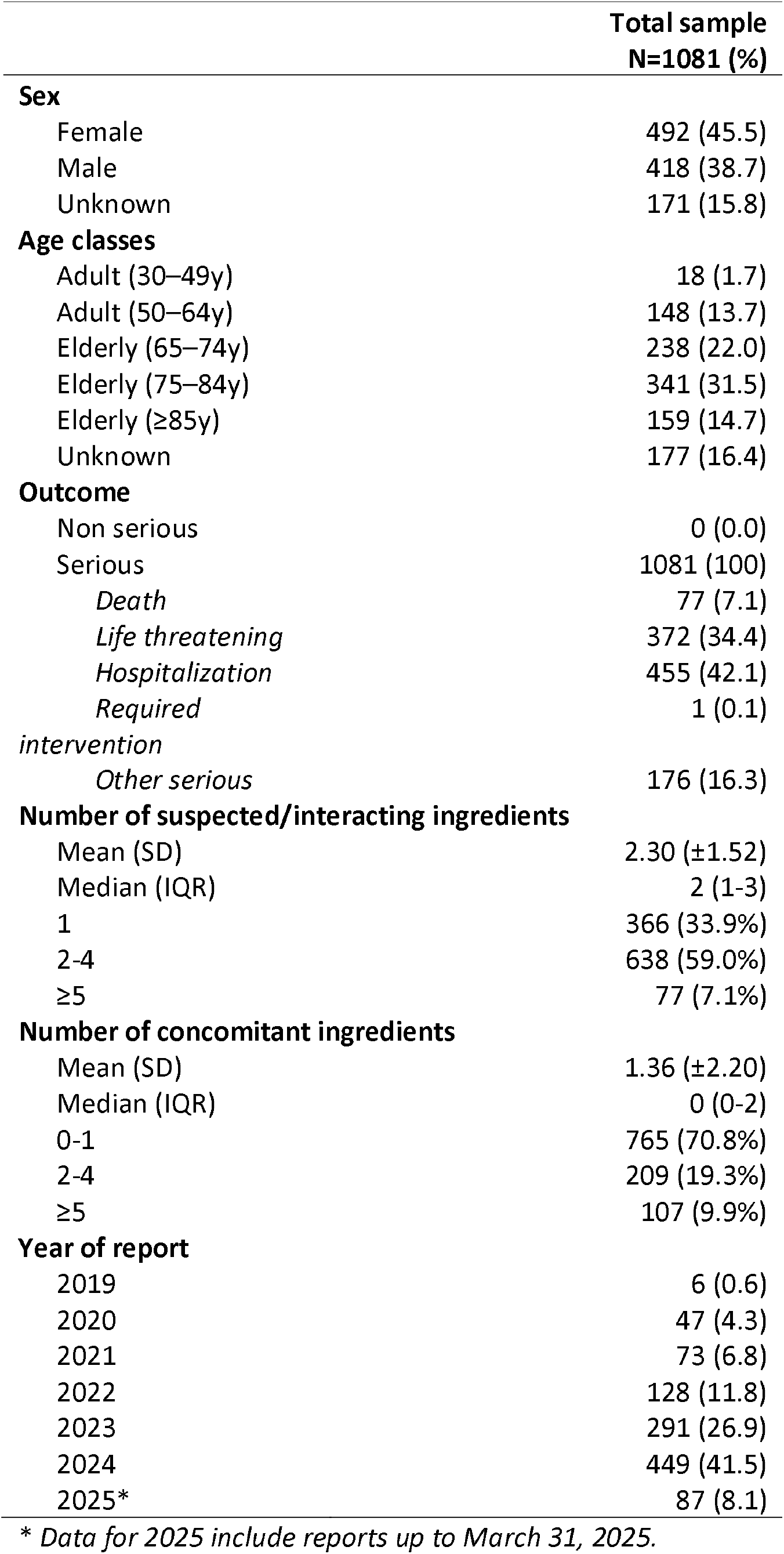
Demographic and clinical characteristics of BRASH syndrome cases reported to FAERS (N=1,081).

Among the 171 Preferred Terms (PTs) reported in association with BRASH syndrome, the most frequently co-reported were overdose (58 cases; 5.37%), toxicity to various agents (44; 4.07%), hyperkalaemia (41; 3.79%), hypotension (40; 3.70%), drug interaction (40; 3.70%), metabolic acidosis (39; 3.61%), cardiac arrest (31; 2.87%), bradycardia (29; 2.68%), and lactic acidosis (28; 2.59%) (**Supplementary Table S1**).

The most frequently reported drugs were metoprolol (591 cases; 54.7%), amlodipine (274; 25.3%), carvedilol (191; 17.7%), furosemide (191; 17.7%), lisinopril (172; 15.9%), spironolactone (143; 13.2%), and verapamil (130; 12.0%) (**Supplementary Table S2**). The most commonly reported indications for suspected or interacting drugs were hypertension (692 cases; 28.8%), atrial fibrillation (191; 7.96%), and heart failure (129; 5.38%). Notably, individual reports frequently included more than one drug suspected or pharmacologically implicated in BRASH syndrome (**Table 1**).

For the purposes of the present analysis, we focused exclusively on drugs pharmacologically implicated in BRASH syndrome, namely atrioventricular nodal blocking agents and drugs involved in the BRASH vicious circle, which are reported in **Table 2**. The complete list of all drugs associated with positive disproportionality signals, including agents without an evident etiopathophysiological role or likely reflecting therapeutic management, is provided in **Supplementary Table S3**.

**Table 2.**
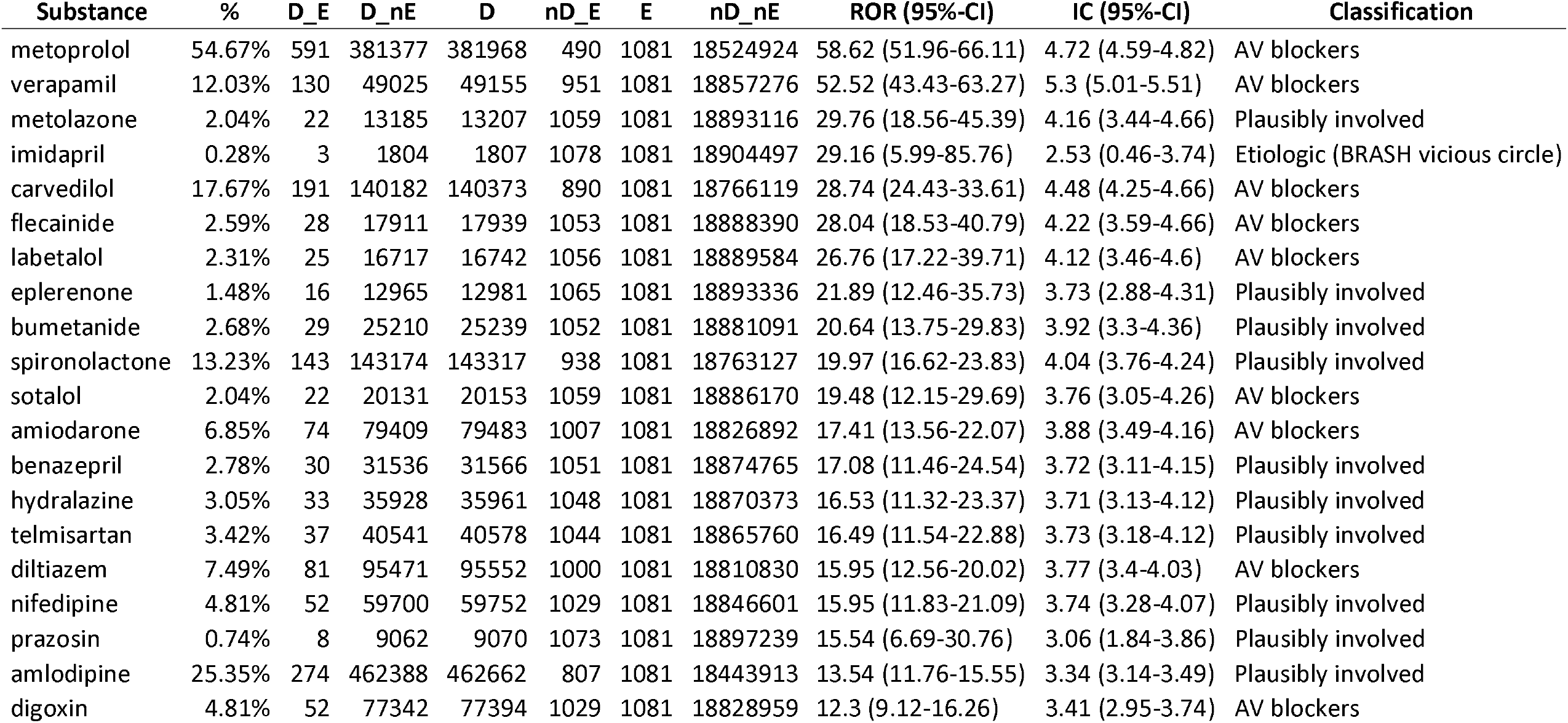
Disproportionality analysis of drug–event pairs reported in association with BRASH syndrome. Only drug–event combinations with at least three reports were included. D_E = number of exposed cases, D_nE = number of non-exposed cases, D = total number of drug reports, E = number of events, nD_E = cases with the event but without the drug of interest, nD_nE = cases without the drug of interest and without the event (reference stratum), ROR (95% CI) = Reporting Odds Ratio with 95% confidence interval, IC (95% CI) = Information Component with 95% confidence interval. “AV blockers” indicates drugs with direct atrioventricular nodal blocking properties; “Plausibly involved” indicates drugs contributing to hyperkalemia, renal impairment, or hypotension within the BRASH pathophysiological cascade.

Disproportionality analyses identified multiple signals of disproportionate reporting between BRASH syndrome and several cardiovascular and renal-active drugs (**Table 2**). The strongest signals were observed for atrioventricular nodal blocking agents, including metoprolol (ROR 58.62, 95% CI 51.96–66.11), verapamil (ROR 52.52, 95% CI 43.43–63.27), and carvedilol (ROR 28.74, 95% CI 24.43–33.61). Significant signals were also detected for drugs plausibly involved in the pathophysiological components of the BRASH vicious circle, such as potassium-sparing diuretics, loop diuretics, and renin–angiotensin system inhibitors.

All reported drug–event pairs met the predefined criteria for signal detection, with at least three reports and lower bounds of the 95% confidence intervals above 1. Bayesian analyses yielded positive Information Component values with lower 95% confidence intervals above zero, supporting the robustness and consistency of the disproportionality findings across different statistical approaches.

Extended disproportionality results, including additional drug–event pairs and context-related medications, are reported in **Supplementary Table S3**.

## DISCUSSION

This study reports the largest series of BRASH syndrome cases to date and represents, to our knowledge, the first pharmacovigilance study specifically dedicated to this clinical entity. BRASH syndrome is a recently described condition, and the progressive increase in reported cases over the past few years likely reflects growing clinical awareness and recognition rather than a true increase in incidence.

The demographic and clinical characteristics observed in this case series indicate that BRASH syndrome predominantly affects elderly patients, with a median age of 74 years, and occurs more frequently in female patients, although sex was not reported in a substantial proportion of cases. These findings suggest that BRASH syndrome primarily manifests in a frail population with multiple comorbidities, particularly cardiovascular disease, including heart failure, hypertension, and atrial fibrillation, and in patients exposed to polypharmacy. The frequent concomitant use of multiple drugs pharmacologically implicated in BRASH syndrome further supports the syndromic and multifactorial nature of this condition.

The predominance of female patients in this study is consistent with the broader pharmacovigilance literature, in which adverse drug reactions are generally reported more frequently in women. However, this finding contrasts with a previously published systematic review of 70 BRASH syndrome cases, where males accounted for the majority of patients (52%) (4,5). This discrepancy may reflect differences in study design, data sources, or reporting patterns, and highlights the need for further investigation in larger and more representative populations.

The advanced age of affected patients is likely explained by the higher prevalence of polypharmacy, chronic cardiovascular and renal disease, and reduced physiological reserve, all of which increase susceptibility to drug–drug interactions and adverse drug events. These factors are central to the pathophysiology of BRASH syndrome, which involves a self-perpetuating cycle of bradycardia, renal dysfunction, and metabolic disturbances.

From a geographical perspective, the marked predominance of reports originating from the United States is expected, given that FAERS is the public pharmacovigilance database of the U.S. Food and Drug Administration (FDA). FAERS collects spontaneous reports submitted by healthcare professionals, patients, and pharmaceutical companies. Moreover, BRASH still remains a significantly underdiagnosed syndromes all over the world affecting real prevalence. Consequently, geographical distributions in FAERS should be interpreted with caution and do not necessarily reflect the true global incidence of the condition.

The severity of the reported events is one of the most concerning aspects. All cases in this series were classified as serious adverse drug reactions, with a high mortality rate (7.1%) or life-threatening conditions (34.4%). These findings suggest that BRASH syndrome represents a critical clinical condition, frequently requiring urgent medical intervention. Notably, the mortality rate observed in this study is higher than that reported in a previous review (5.7%) (2). In addition, the most frequently co-reported Preferred Terms—such as hypotension, bradycardia, metabolic acidosis, and cardiac arrest—further underscore the severity and multisystem involvement characterizing BRASH syndrome.

The pharmacological patterns observed in this study are highly consistent with the multifactorial pathophysiology of BRASH syndrome, in which converging drug effects on cardiac conduction, renal function, and electrolyte balance interact within a self-perpetuating cycle. The most frequently reproted drugs associated with BRASH syndrome included atrioventricular nodal blocking agents, such as beta-blockers and non-dihydropyridine calcium channel blockers, as well as antihypertensive agents and diuretics that may contribute to renal dysfunction and hyperkalaemia.

In addition, antiarrhythmics, such as metoprolol, carvedilol, verapamil, and flecainide, as well as antihypertensives like imidapril and labetalol, diuretics as metolazone and potassium sparing agents are often associated with BRASH

**Metoprolol** is the most widely prescribed beta-blocker and belongs to the Vaughan-Williams class II antiarrhythmics (6). As a cardioselective beta-blocker, it is a preferred agent for heart failure with reduced ejection fraction (HFrEF) and is widely used to treat hypertension, angina, and for rate control in atrial fibrillation and ventricular arrhythmias (7,8). Metoprolol is eliminated almost exclusively via hepatic metabolism, primarily by cytochrome P450 (CYP) 2D6. Consequently, factors such as age, ethnicity, concomitant use of CYP2D6 inhibitors and CYP polymorphisms may alter its bioavailability (9).

**Carvedilol** is another frequently used agent, functioning as a non-selective beta-blocker with alpha-1 blocking activity. Similar to other beta-blockers, its plasma levels are influenced by the hepatic first-pass effect (10). It is widely prescribed for the management of heart failure (8).

**Labetalol** is a combined alpha- and beta-adenoceptor blocking agent for oral and intravenous use in the treatment of hypertension. It is a nonselective competitive antagonist at beta-adrenoceptors and a competitive antagonist of postsynaptic alpha_1_-adrenoceptors. Labetalol is more potent at beta than at alpha_1_ adrenoceptors (11). It reduces blood pressure to a similar extent, and in a similar proportion of patients, as ‘pure’ beta-blockers such as propranolol, pure alpha-blockers such as prazosin, calcium antagonists (nifedipine, verapamil), and centrally acting drugs (clonidine and methyldopa). Labetalol is very effective in hypertensive pregnant women and in hypertensive crises (12).

**Verapamil** is a non-dihydropyridine calcium channel blocker belonging to the Vaughan-Williams class IV antiarrhythmics. It undergoes extensive hepatic extraction, resulting in low systemic bioavailability following oral administration (13). Verapamil is largely metabolized by CYP3A4 and acts both as a substrate and an inhibitor of this enzyme (14). It is widely indicated for hypertension, angina, rate-control in atrial fibrillation and flutter, and complex arrhythmias such as paroxysmal supraventricular tachycardia and fascicular ventricular tachycardia (15,16).

Both metoprolol and verapamil exhibit negative chronotropic and inotropic effects. During acute illness, the combined reduction in heart rate and contractility can cause renal hypoperfusion, fluid retention, and hyperkalemia; this leads to drug accumulation and atrioventricular (AV) block, thereby triggering and amplifying the BRASH syndrome spiral.

**Flecainide** is a class Ic antiarrhythmic that reduces myocardial contractility primarily by inhibiting fast sodium channels and L-type calcium channels, leading to decreased intracellular calcium availability during excitation-contraction coupling (17). This results in a dose-dependent reduction in isometric contractility and reduced muscle fiber recruitment. This negative inotropic effect is clinically significant and may be more pronounced at higher plasma concentrations or in patients with pre-existing left ventricular dysfunction. The Cardiac Arrhythmia Suppression Trial demonstrated that in patients with structural heart disease or recent myocardial infarction, flecainide led to a significant increase in mortality compared to placebo (18). In older patients, reduced cardiac function combined with acute intercurrent events (e.g., dehydration, infection) can trigger the lethal BRASH syndrome spiral.

**Digoxin** is a cardiac glycoside with a negative chronotropic effect and a narrow therapeutic index. It is eliminated primarily through kidney, and therefore serum digoxin concentrations are highly sensitive to changes in renal function. Conditions such as dehydration or acute kidney injury may lead to reduced clearance and increased serum levels, thereby enhancing its electrophysiological effects and predisposing to bradyarrhythmias (19–21).

**Amiodarone** was approved by FDA for the first time on December 1985 for treatment of life-threatening ventricular tachyarrhythmias. It is effective not only in these arrhythmias but also in less severe ventricular arrhythmias and many supraventricular arrhythmias, including atrial fibrillation and reentrant tachyarrhythmias involving accessory pathways (22). It is classified electrophysiologically as a Type III antiarrhythmic. Its negative inotropic effect is usually negligible. Amiodarone is poorly bioavailable (20-80%) and undergoes extensive enterohepatic circulation before entry into a central compartment. The principal metabolite, mono-n-desethyl amiodarone is also an antiarrhythmic. From this central compartment, it undergoes extensive tissue distribution (exceptionally high tissue/plasma partition coefficients). The distribution half-life of amiodarone out of the central compartment to peripheral and deep tissue compartments (t_1/2,α_) may be as short as 4 hours. The terminal half-life (t_1/2,β_) is both long and variable (9-77 days) secondary to the slow mobilization of the lipophilic medication out of (primarily) adipocytes (23,24). However, the drug has several side effects, including thyroid abnormalities, pulmonary fibrosis, and transaminitis, for which routine monitoring is recommended. It also interacts with several medications, such as warfarin, simvastatin, and atorvastatin, and many HIV antiretroviral medications (25). Its negative inotropic effect and its atrioventricular nodal blocking action, associated to its own pharmacokinetic properties, may be involved in developing of BRASH syndrome.

Importantly, the contribution of atrioventricular nodal blocking agents to BRASH syndrome should not be interpreted solely as the consequence of increased plasma drug concentrations due to reduced renal clearance. Although acute kidney injury may lead to some degree of drug accumulation, synergistic electrophysiological mechanisms likely play a major role. Hyperkalaemia reduces the myocardial resting membrane potential, slows phase 0 depolarization in nodal tissue, and decreases atrioventricular conduction velocity. Consequently, even modest increases in serum potassium levels may markedly potentiate the negative chronotropic and negative dromotropic effects of beta-blockers and non-dihydropyridine calcium channel blockers at the atrioventricular node.

In addition, acute renal dysfunction is frequently accompanied by metabolic acidosis, uremia, and alterations in ionized calcium, all of which further increase myocardial sensitivity and reduce electrophysiological reserve. These metabolic disturbances may amplify the functional effects of AV nodal blocking agents, leading to profound bradycardia and conduction disturbances through synergistic rather than purely cumulative mechanisms. Emerging evidence suggests that hypocalcemia may precipitate a clinically indistinguishable ‘BRASH-like’ syndrome. In these cases, low ionized calcium exacerbates the negative inotropic and dromotropic effects of AV-blockers, leading to shock and bradycardia even in the absence of severe hyperkalemia (26). Additional signals involved drugs contributing to the renal and electrolyte components of the syndrome.

**Imidapril** is an angiotensin-converting enzyme (ACE) inhibitor used to treat hypertension, heart failure, and diabetic nephropathy (21). By inhibiting ACE during acute illnesses involving dehydration or sepsis, imidapril may aggravate renal failure and cause potassium retention; this can lead to AV block, initiating the BRASH syndrome cascade.

**Metolazone** is an oral quinazoline diuretic with thiazide-like properties, indicated for salt and water retention associated with congestive heart failure and renal disease. It acts primarily by inhibiting sodium reabsorption at the cortical diluting site and the proximal convoluted tubule, increasing sodium and chloride excretion (27). Metolazone is notable for its efficacy in patients with severe renal failure (GFR < 20 ml/min). However, due to its potent diuretic effect, it may cause electrolyte disturbances and worsen renal function; this can potentiate concomitant diuretics in patients with limited renal reserve, promoting hyperkalemia and reducing the clearance of AV nodal blockers, thus precipitating BRASH syndrome (28).

**Eplerenone** and **spironolactone** are potassium-sparing diuretics used for heart failure and hypertension:

- **Spironolactone** is a mineralocorticoid receptor antagonist structurally similar to progesterone and is associated with progestogenic and antiandrogenic effects (29). It undergoes rapid, extensive metabolism into three active metabolites with prolonged half-lives (up to 16.5 hours).
- **Eplerenone** is a derivative with lower affinity for progesterone and androgen receptors. It also undergoes extensive metabolism, but its metabolites are inactive, and it has a shorter elimination half-life (up to 6 hours) (30).

Both agents are associated with dose-related increases in serum potassium levels. Patients with underlying renal dysfunction or heart disease are at the greatest risk of hyperkalemia (31,32). The extended half-life of spironolactone’s active metabolites may further increase the risk of hyperkalemia and its complications (33). Hyperkalemia can impair electrical impulse conduction at the AV node, leading to severe bradycardia or AV block, potentially initiating or worsening BRASH syndrome.

### Strengths and limitations

This study has several limitations inherent to pharmacovigilance analyses based on spontaneous reporting systems. Adverse event reports are often incomplete, particularly with regard to drug exposure details such as treatment duration, dosage, and the temporal relationship between drug intake and event onset. In addition, spontaneous reporting systems are subject to reporting and selection biases, as more severe or clinically relevant cases are more likely to be reported, potentially leading to an overrepresentation of serious outcomes.

Another limitation of this study is case misclassification bias, as case identification relied exclusively on reports explicitly coded as “*BRASH syndrome*”, potentially resulting in under-ascertainment of clinically compatible cases reported under related individual manifestations. Finally, given the relatively recent recognition of BRASH syndrome as a distinct clinical entity, increased awareness among clinicians may have contributed to stimulated reporting over time, introducing temporal reporting trends that may not reflect true incidence. (34,35).

Despite these limitations, this study has several important strengths. To our knowledge, it represents the first large-scale pharmacovigilance analysis to investigate BRASH syndrome using a substantial number of reported cases, enabling a comprehensive evaluation of clinical outcomes at the population level. Moreover, this analysis extends beyond atrioventricular nodal blocking agents to examine the role of other commonly prescribed medications, including antihypertensive agents, potassium-sparing drugs, and diuretics. This comprehensive pharmacological assessment supports the interpretation of BRASH syndrome as a complex clinical syndrome driven by polypharmacy and drug–drug interactions rather than as a single, isolated adverse drug reaction.

## CONCLUSION

Overall, these findings support the concept that BRASH syndrome should not be interpreted as a single-drug adverse reaction, but rather as the result of converging pharmacological mechanisms occurring in vulnerable patients.

Numerous medications with distinct mechanisms can lead to BRASH syndrome. Antiarrhythmic drugs may impair atrioventricular and intraventricular conduction, resulting in negative chronotropic and inotropic effects; this causes renal dysfunction, ionic imbalance, and fluid retention, which increases plasma drug levels and fuels the negative spiral of the syndrome. Concurrently, antihypertensives may cause kidney injury and hyperkalemia, increasing plasma concentrations of other drugs, such as antiarrhythmics and diuretics, thereby starting or amplifying BRASH syndrome.

Generally, patients requiring these heart-active medications are frail, have multiple comorbidities, are exposed to polypharmacy, and often suffer from co-existing kidney disease or chronic cardio-renal syndrome. Intercurrent events such as sepsis, infection, or dehydration can lead to increased plasma levels of the aforementioned medications, worsening both heart and renal function. Accordingly, a scoring system applied before the prescription of cardioactive medications in patients with specific clinical conditions or impaired heart, kidney or renal function could be useful to prevent BRASH syndrome.

## Data Availability

All data are extracted by FAERS database. Analysis of data are available asking to first two authors.

