## supplementary for "BRASH syndrome as a clinical syndrome driven by polypharmacy: a pharmacovigilance study of 1,081 cases from FAERS"

**Table S1.** The 20 most frequently reported PTs.

1. brash syndrome (100%) [1081]

2. overdose (5.37%) [58]

3. toxicity to various agents (4.07%) [44]

4. hyperkalaemia (3.79%) [41]

5. hypotension (3.7%) [40]

6. drug interaction (3.7%) [40]

7. metabolic acidosis (3.61%) [39]

8. cardiac arrest (2.87%) [31]

9. bradycardia (2.68%) [29]

10. lactic acidosis (2.59%) [28]

11. off label use (2.41%) [26]

12. atrial fibrillation (2.41%) [26]

13. respiratory failure (2.22%) [24]

14. encephalopathy (1.85%) [20]

15. cardiogenic shock (1.76%) [19]

16. acute kidney injury (1.67%) [18]

17. multiple organ dysfunction syndrome (1.67%) [18]

18. diarrhoea (1.57%) [17]

19. hypovolaemia (1.57%) [17]

20. hyponatraemia (1.57%) [17]

**Table S2.** The 20 most frequently reported drugs.

| **Substance** | **N_cases** | **%** |
| --- | --- | --- |
| metoprolol | 591 | 54.7% |
| amlodipine | 274 | 25.3% |
| carvedilol | 191 | 17.7% |
| furosemide | 191 | 17.7% |
| lisinopril | 172 | 15.9% |
| spironolactone | 143 | 13.2% |
| verapamil | 130 | 12.0% |
| losartan | 122 | 11.3% |
| metformin | 102 | 9.4% |
| valsartan | 93 | 8.6% |
| apixaban | 81 | 7.5% |
| diltiazem | 81 | 7.5% |
| amiodarone | 74 | 6.8% |
| atorvastatin | 72 | 6.7% |
| bisoprolol | 70 | 6.5% |
| acetylsalicylic acid | 68 | 6.3% |
| insulin | 53 | 4.9% |
| digoxin | 52 | 4.8% |
| nifedipine | 52 | 4.8% |
| levothyroxine | 44 | 4.1% |

**Table S3.**

| **substance** | **Prevalence in BRASH cases** | **D_E** | **D_nE** | **D** | **nD_E** | **E** | **nD_nE** | **ROR (95%-CI)** | **IC (95%-CI)** | **Classification** |
| --- | --- | --- | --- | --- | --- | --- | --- | --- | --- | --- |
| metoprolol | 54.67% | 591 | 381377 | 381968 | 490 | 1081 | 18524924 | 58.62 (51.96-66.11) | 4.72 (4.59-4.82) | AV blockers |
| glucagon | 1.30% | 14 | 4414 | 4428 | 1067 | 1081 | 18901887 | 56.19 (30.58-94.92) | 4.26 (3.36-4.88) | Likely therapeutic drugs |
| verapamil | 12.03% | 130 | 49025 | 49155 | 951 | 1081 | 18857276 | 52.52 (43.43-63.27) | 5.3 (5.01-5.51) | AV blockers |
| vasopressin | 1.11% | 12 | 4154 | 4166 | 1069 | 1081 | 18902147 | 51.11 (26.28-89.64) | 4.08 (3.1-4.74) | Likely therapeutic drugs |
| dopamine | 1.30% | 14 | 6215 | 6229 | 1067 | 1081 | 18900086 | 39.9 (21.72-67.25) | 4.08 (3.17-4.7) | Likely therapeutic drugs |
| metolazone | 2.04% | 22 | 13185 | 13207 | 1059 | 1081 | 18893116 | 29.76 (18.56-45.39) | 4.16 (3.44-4.66) | Drugs plausibly involved in the BRASH pathophysiological cascade |
| imidapril | 0.28% | 3 | 1804 | 1807 | 1078 | 1081 | 18904497 | 29.16 (5.99-85.76) | 2.53 (0.46-3.74) | Drugs plausibly involved in the BRASH pathophysiological cascade |
| carvedilol | 17.67% | 191 | 140182 | 140373 | 890 | 1081 | 18766119 | 28.74 (24.43-33.61) | 4.48 (4.25-4.66) | AV blockers |
| flecainide | 2.59% | 28 | 17911 | 17939 | 1053 | 1081 | 18888390 | 28.04 (18.53-40.79) | 4.22 (3.59-4.66) | AV blockers |
| labetalol | 2.31% | 25 | 16717 | 16742 | 1056 | 1081 | 18889584 | 26.76 (17.22-39.71) | 4.12 (3.46-4.6) | AV blockers |
| norepinephrine | 1.85% | 20 | 15026 | 15046 | 1061 | 1081 | 18891275 | 23.69 (14.41-36.8) | 3.91 (3.16-4.43) | Likely therapeutic drugs |
| dobutamine | 0.74% | 8 | 6213 | 6221 | 1073 | 1081 | 18900088 | 22.67 (9.76-44.88) | 3.31 (2.09-4.11) | Likely therapeutic drugs |
| polaprezinc | 0.28% | 3 | 2402 | 2405 | 1078 | 1081 | 18903899 | 21.9 (4.5-64.24) | 2.45 (0.38-3.66) | Associated drug, not etiologic |
| eplerenone | 1.48% | 16 | 12965 | 12981 | 1065 | 1081 | 18893336 | 21.89 (12.46-35.73) | 3.73 (2.88-4.31) | Drugs plausibly involved in the BRASH pathophysiological cascade |
| atropine | 3.33% | 36 | 30183 | 30219 | 1045 | 1081 | 18876118 | 21.54 (14.99-30.02) | 4.03 (3.47-4.42) | Likely therapeutic drugs |
| bumetanide | 2.68% | 29 | 25210 | 25239 | 1052 | 1081 | 18881091 | 20.64 (13.75-29.83) | 3.92 (3.3-4.36) | Drugs plausibly involved in the BRASH pathophysiological cascade |
| spironolactone | 13.23% | 143 | 143174 | 143317 | 938 | 1081 | 18763127 | 19.97 (16.62-23.83) | 4.04 (3.76-4.24) | Drugs plausibly involved in the BRASH pathophysiological cascade |
| sotalol | 2.04% | 22 | 20131 | 20153 | 1059 | 1081 | 18886170 | 19.48 (12.15-29.69) | 3.76 (3.05-4.26) | AV blockers |
| amiodarone | 6.85% | 74 | 79409 | 79483 | 1007 | 1081 | 18826892 | 17.41 (13.56-22.07) | 3.88 (3.49-4.16) | AV blockers |
| benazepril | 2.78% | 30 | 31536 | 31566 | 1051 | 1081 | 18874765 | 17.08 (11.46-24.54) | 3.72 (3.11-4.15) | Drugs plausibly involved in the BRASH pathophysiological cascade |
| hydralazine | 3.05% | 33 | 35928 | 35961 | 1048 | 1081 | 18870373 | 16.53 (11.32-23.37) | 3.71 (3.13-4.12) | Drugs plausibly involved in the BRASH pathophysiological cascade |
| telmisartan | 3.42% | 37 | 40541 | 40578 | 1044 | 1081 | 18865760 | 16.49 (11.54-22.88) | 3.73 (3.18-4.12) | Drugs plausibly involved in the BRASH pathophysiological cascade |
| diltiazem | 7.49% | 81 | 95471 | 95552 | 1000 | 1081 | 18810830 | 15.95 (12.56-20.02) | 3.77 (3.4-4.03) | AV blockers |
| nifedipine | 4.81% | 52 | 59700 | 59752 | 1029 | 1081 | 18846601 | 15.95 (11.83-21.09) | 3.74 (3.28-4.07) | Drugs plausibly involved in the BRASH pathophysiological cascade |
| prazosin | 0.74% | 8 | 9062 | 9070 | 1073 | 1081 | 18897239 | 15.54 (6.69-30.76) | 3.06 (1.84-3.86) | Drugs plausibly involved in the BRASH pathophysiological cascade |
| amlodipine | 25.35% | 274 | 462388 | 462662 | 807 | 1081 | 18443913 | 13.54 (11.76-15.55) | 3.34 (3.14-3.49) | Drugs plausibly involved in the BRASH pathophysiological cascade |
| digoxin | 4.81% | 52 | 77342 | 77394 | 1029 | 1081 | 18828959 | 12.3 (9.12-16.26) | 3.41 (2.95-3.74) | AV blockers |
| amiloride | 0.56% | 6 | 8692 | 8698 | 1075 | 1081 | 18897609 | 12.13 (4.44-26.5) | 2.7 (1.28-3.61) | Drugs plausibly involved in the BRASH pathophysiological cascade |
| azilsartan | 0.37% | 4 | 5995 | 5999 | 1077 | 1081 | 18900306 | 11.7 (3.18-30.07) | 2.41 (0.65-3.49) | Drugs plausibly involved in the BRASH pathophysiological cascade |
| losartan | 11.29% | 122 | 217722 | 217844 | 959 | 1081 | 18688579 | 10.91 (8.96-13.2) | 3.24 (2.94-3.45) | Drugs plausibly involved in the BRASH pathophysiological cascade |
| lisinopril | 15.91% | 172 | 327229 | 327401 | 909 | 1081 | 18579072 | 10.74 (9.07-12.65) | 3.16 (2.91-3.34) | Drugs plausibly involved in the BRASH pathophysiological cascade |
| epinephrine | 3.42% | 37 | 68380 | 68417 | 1044 | 1081 | 18837921 | 9.76 (6.83-13.54) | 3.08 (2.53-3.47) | Likely therapeutic drugs |
| furosemide | 17.67% | 191 | 411725 | 411916 | 890 | 1081 | 18494576 | 9.63 (8.2-11.28) | 2.99 (2.75-3.16) | Drugs plausibly involved in the BRASH pathophysiological cascade |
| empagliflozin | 2.96% | 32 | 59939 | 59971 | 1049 | 1081 | 18846362 | 9.59 (6.52-13.62) | 3.04 (2.45-3.46) | Associated drug, not etiologic |
| ranolazine | 0.93% | 10 | 18869 | 18879 | 1071 | 1081 | 18887432 | 9.34 (4.46-17.25) | 2.73 (1.65-3.45) | Drugs plausibly involved in the BRASH pathophysiological cascade |
| isosorbide dinitrate | 0.65% | 7 | 13478 | 13485 | 1074 | 1081 | 18892823 | 9.13 (3.66-18.89) | 2.56 (1.25-3.41) | Drugs plausibly involved in the BRASH pathophysiological cascade |
| bisoprolol | 6.48% | 70 | 161606 | 161676 | 1011 | 1081 | 18744695 | 8.03 (6.21-10.23) | 2.85 (2.45-3.14) | AV blockers |
| valsartan | 8.60% | 93 | 263755 | 263848 | 988 | 1081 | 18642546 | 6.65 (5.31-8.23) | 2.58 (2.24-2.83) | Drugs plausibly involved in the BRASH pathophysiological cascade |
| apixaban | 7.49% | 81 | 243859 | 243940 | 1000 | 1081 | 18662442 | 6.19 (4.88-7.77) | 2.49 (2.12-2.76) | Associated drug, not etiologic |
| dapagliflozin | 1.57% | 17 | 50421 | 50438 | 1064 | 1081 | 18855880 | 5.97 (3.46-9.61) | 2.37 (1.55-2.93) | Associated drug, not etiologic |
| torasemide | 1.67% | 18 | 53759 | 53777 | 1063 | 1081 | 18852542 | 5.93 (3.5-9.43) | 2.37 (1.57-2.92) | Drugs plausibly involved in the BRASH pathophysiological cascade |
| indapamide | 0.83% | 9 | 27506 | 27515 | 1072 | 1081 | 18878795 | 5.76 (2.62-10.97) | 2.19 (1.05-2.95) | Drugs plausibly involved in the BRASH pathophysiological cascade |
| atenolol | 3.98% | 43 | 137718 | 137761 | 1038 | 1081 | 18768583 | 5.64 (4.06-7.65) | 2.37 (1.86-2.73) | AV blockers |
| olmesartan | 1.94% | 21 | 66184 | 66205 | 1060 | 1081 | 18840117 | 5.63 (3.47-8.66) | 2.32 (1.59-2.83) | Drugs plausibly involved in the BRASH pathophysiological cascade |
| clonidine | 1.76% | 19 | 64832 | 64851 | 1062 | 1081 | 18841469 | 5.2 (3.11-8.15) | 2.21 (1.44-2.75) | Drugs plausibly involved in the BRASH pathophysiological cascade |
| perindopril | 1.39% | 15 | 52924 | 52939 | 1066 | 1081 | 18853377 | 5.01 (2.79-8.3) | 2.13 (1.26-2.73) | Drugs plausibly involved in the BRASH pathophysiological cascade |
| glimepiride | 1.57% | 17 | 63861 | 63878 | 1064 | 1081 | 18842440 | 4.71 (2.73-7.58) | 2.07 (1.25-2.64) | Associated drug, not etiologic |
| sacubitril | 2.68% | 29 | 117866 | 117895 | 1052 | 1081 | 18788435 | 4.39 (2.92-6.34) | 2.02 (1.4-2.46) | Associated drug, not etiologic |
| pravastatin | 2.13% | 23 | 98635 | 98658 | 1058 | 1081 | 18807666 | 4.14 (2.61-6.25) | 1.93 (1.23-2.42) | Associated drug, not etiologic |
| metformin | 9.44% | 102 | 500337 | 500439 | 979 | 1081 | 18405964 | 3.83 (3.09-4.7) | 1.81 (1.48-2.05) | Associated drug, not etiologic |
| enalapril | 1.39% | 15 | 70951 | 70966 | 1066 | 1081 | 18835350 | 3.73 (2.08-6.18) | 1.76 (0.89-2.36) | Drugs plausibly involved in the BRASH pathophysiological cascade |
| propranolol | 1.20% | 13 | 73527 | 73540 | 1068 | 1081 | 18832774 | 3.11 (1.65-5.35) | 1.52 (0.58-2.16) | AV blockers |
| ramipril | 2.13% | 23 | 146382 | 146405 | 1058 | 1081 | 18759919 | 2.78 (1.75-4.2) | 1.4 (0.7-1.89) | Drugs plausibly involved in the BRASH pathophysiological cascade |
| atorvastatin | 6.66% | 72 | 498793 | 498865 | 1009 | 1081 | 18407508 | 2.63 (2.04-3.34) | 1.32 (0.93-1.6) | Associated drug, not etiologic |
| rosuvastatin | 2.68% | 29 | 208944 | 208973 | 1052 | 1081 | 18697357 | 2.46 (1.64-3.56) | 1.24 (0.62-1.68) | Associated drug, not etiologic |
| insulin | 4.90% | 53 | 516700 | 516753 | 1028 | 1081 | 18389601 | 1.83 (1.36-2.41) | 0.83 (0.37-1.15) | Likely therapeutic drugs |
